# Classifying Refugee Status Using Common Features in EMR

**DOI:** 10.1101/2021.08.17.21262048

**Authors:** Malia Morrison, Vanessa Nobles, Crista E. Johnson-Agbakwu, Celeste Bailey, Li Liu

**Affiliations:** College of Health Solutions, Arizona State University, Phoenix, AZ, 85004, USA; Department of Obstetrics, Gynecology and Women’s Health, Valleywise Health, Phoenix, AZ, 85008, USA; Creighton University School of Medicine -Phoenix Campus, Phoenix, AZ, 85008, USA; District Medical Group, Mesa, AZ, 85201, USA; Southwest Interdisciplinary Research Center, Watts College of Public Service and Community Solutions, Arizona State University, Tempe, AZ; Biodesign Institute, Arizona State University, Tempe, AZ, 85281, USA; Department of Neurology, Mayo Clinic, Scottsdale, AZ 85259, USA

**Keywords:** refugee health, machine learning, health disparity, health informatics

## Abstract

**Objective:** Automated and accurate identification of refugees in healthcare databases is a critical first step to investigate healthcare needs of this vulnerable population and improve health disparities. This study developed a machine-learning method, named refugee identification system (RIS) that uses features commonly collected in healthcare databases to classify refugees and non-refugees.

**Materials and Methods:** We compiled a curated data set consisting of 103 refugees and 930 non-refugees in Arizona. For each person in the curated data set, we collected age, primary language, and noise-masked home address. We supplemented de-identified individual-level data with state-level refugee resettlement statistics and world language statistics, then performed feature engineering to convert primary language and masked address into quantitative features. Finally, we built a random forest model to classify refugee status.

**Results:** Evaluated on holdout testing data, RIS achieved a high classification accuracy of 0.97, specificity of 0.99, sensitivity of 0.85, positive predictive value of 0.88, and negative predictive value of 0.98. The receiver operating characteristic curve had an area under the curve value of 0.98. The source code is available at GitHub (https://github.com/liliulab/ris).

**Discussion and Conclusion:** RIS is an automated, accurate, and scalable method to predict refugee status. It uses only de-identified information to protect patient privacy. The computational framework is adaptable to address similar challenges in other States. Its application enables large-scale investigation of refugee healthcare needs and improvement of health disparities.

## INTRODUCTION

According to the Immigration and Nationality Act of the United States, a refugee is a person who has fled their country due to persecution or fear of persecution and is unwilling or unable to return, due to one’s race, religion, nationality, membership in a particular social group, or political opinion ^[1]^. In the last two decades, there have been over 1 million refugees admitted to the United States^[2]^. Approximately 5% of those refugees have been resettled in the state of Arizona^[3]^.

Refugees often come from war-torn regions where access to healthcare is limited^[4]^. They face unique physical and mental health concerns after surviving civil conflicts, gender-based violence, and other human rights atrocities, as well as protracted time in precarious refugee camp settings. Once they have been resettled in host countries, refugees continue to face further challenges accessing and navigating the healthcare system amidst language, literacy, and communication barriers alongside other social determinants of health^[5]^.

To analyze refugee-specific health outcomes and improve health disparities, identifying these individuals in healthcare networks is a prerequisite. Unfortunately, refugee status is not stored in electronic medical records (EMR), electronic health records (EHR), insurance claims, and other healthcare databases^[6]^. Scientists have experimented automated approaches, such as searching keywords in unstructured text of EMRs^[7]^. However, these approaches can identify only predefined terms and follow rigid rules that are not flexible to accommodate general settings. Consequently, practitioners still rely on manual reviews to distinguish refugees from non-refugees, which is inefficient, unscalable, and difficult to share^[8]^. Intelligent computational methods that can be efficiently and effectively applied across multiple healthcare databases to identify refugees are needed urgently.

To explore features informative of refugee status in healthcare databases, we examined the EMR (Epic) system at Valleywise Health (VH) in Maricopa County, Arizona, that is the only public, teaching, safety net health care system in the state of Arizona with the mission to care for marginalized and underserved populations. We manually curated the refugee status of 1,033 individuals (103 refugees and 930 non-refugees) and found several features informative of an individual’s refugee status, such as primary language, citizenship, and past visits to Refugee Clinics. However, not all these features are commonly recorded in healthcare databases at other organizations. For example, citizenship is seldom specified, and visits to Refugee Clinics are not shared between systems. A classification model using these features will have limited applicability and generalizability at the state-level or nation-level. Meanwhile, more than 7,000 languages are spoken worldwide^[9]^. Even after eliminating extremely rare languages, there remain 200 languages that are the native tongue of over 88% of the global population. Furthermore, refugee resettlement is highly dynamic with home countries varying significantly over time^[10]^. Such high diversity and complexity challenge feature encoding as it is not feasible to have all languages and citizenships represented in the curated data set.

In this study, we present a novel machine-learning method, named Refugee Identification System (RIS). RIS distinguishes refugees and non-refugees using information readily available in EMR and EHR. To account for languages not represented in curated data, we innovatively used state-level refugee statistics and world language statistics as supplementary information for feature engineering. Via automated and accurate classifications, RIS helps reduce manual review effort. Accurate and efficient identification of refugees allows refugee-specific health data to be extracted, updated, and analyzed in a timely manner, ensuring this vulnerable population receives the specialized care they require. The cross-platform implementation of this method is freely available at GitHub (https://github.com/liliulab/ris).

## MATERIALS AND METHODS

### Dataset

The dataset consisted of 1,033 deidentified individuals randomly selected from the VH EMR database (Epic) who visited the VH Women’s Health Center between May 2020 and December 2020. Via manual curation by team experts, we confirmed the refugee status of these individuals including 103 refugees and 930 non-refugees. The study was approved by the VH Institutional Review Board (#2011-007).

For each individual, we extracted primary language, home address, and age from the EMR as raw features. Because these raw features are not quantitative or exhaustive of all potential values, we performed feature engineering to: (1) convert languages into quantitative scores based on the relative frequency of which they are spoken by refugees and non-refugees, and (2) convert addresses into physical distances from residential clusters.

### Feature engineering of languages

The Arizona Department of Economic Security releases an annual refugee arrival report (AZRAR) of the total number of refugees from each country who have been resettled in Arizona since 1975^[11]^. To better reflect the current landscape of refugee home countries, we removed records from the AZRAR before the year 2000. A total of *N* = 56,907 refugees from 96 countries have been resettled in Arizona since 2000^[3]^. For each country *c*, we denoted the number of refugees resettled in Arizona from that country as *n*_*c*_, and computed the fraction,

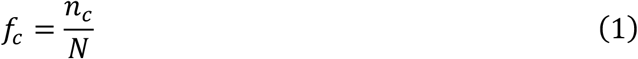

The country with the highest fraction was Iraq (*f*_*c*_ = 0.18). Six countries tied with the lowest fraction of *f*_*c*_ = 0.0, including Djibouti, Ghana, Hungary, Latvia, Lithuania, and Poland.

We then used the CIA’s World Factbook to retrieve languages spoken in each country^[12]^. Given a country *c*, if the Factbook provided the percentage of the population speaking a given language *l*, we retrieved the percentage directly and denoted it as *p*_*l,c*_. If the Factbook simply listed languages spoken in country *c* in descending order from most to least commonly spoken, we converted the ranked orders to an artificial percentage based on Roulette wheel selection probability^[13]^,

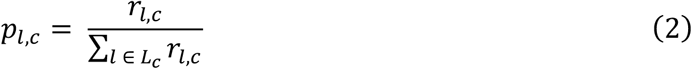

where *r*_*l,c*_ is the language’s rank within country *c*, and *L*_*c*_ is all languages spoken in country *c*. We then aggregated *p*_*l,c*_ across all countries *C* to produce a weighted score *s*_*l*_ adjusting for country-specific refugee fraction *f*_*c*_,

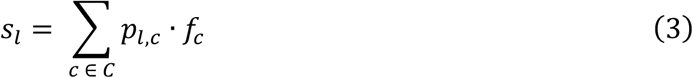

Lastly, we created a ranked list of languages based on *s*_*l*_, in which the language with the highest *s*_*l*_ value received rank 1. In case of ties where multiple languages had the same weighted scores, we found their lowest ranking and assigned this ranking to all languages in the tie.

### Feature engineering of home addresses

We converted all addresses into latitude and longitude coordinates using the Python GeoPy client. To protect patient privacy, we converted home addresses to de-identified information by adding noise ranging from 0.347 to 0.542 miles to the latitude and longitude coordinates. Via hierarchical clustering based on Euclidean distances and Ward’s minimum variance linkage, we organized residential addresses into a tree structure (i.e., dendrogram) for refugees and non-refugees separately. By examining branch lengths of the dendrograms, we determined the cutoff to partition the trees into disjoint clusters (cutoff=0.35 and 0.8 for the refugee tree and the non-refugee tree, respectively). For each location cluster, we computed the centroid and its corresponding latitude and longitude coordinates. Given an individual’s residential address plus the buffer, we calculated its distance to each of the centroids and used these distances as quantitative features.

### Predictive model of refugee status

The RIS is a random forests model to classify refugees and non-refugees.

We split the curated data into a training set consisting of 723 randomly selected individuals (70%) and a testing set consisting of the remaining 310 individuals (30%). For cross-validations, we further split the training set in three folds, used two folds for training and the remaining fold for validation, and repeated three times. The prevalence rate of refugees was approximately 10%, making the data set highly imbalanced. To correct this bias, we applied the Synthetic Minority Oversampling TEchnique (SMOTE) to the training data^[14]^. We did not apply SMOTE to the validation and testing data, so that the model performance reflected the overabundance of the non-refugee population in real world applications.

To determine the hyperparameters of the random forest model, we performed a grid search with the configurations specified in **Table 1**.

**Table 1.** Configuration of grid search for optimal hyperparameters.

| Hyperparameter | Min | Max | Interval |
| --- | --- | --- | --- |
| Max features | 15 | 22 | 1 |
| Min samples per leaf | 10 | 50 | 5 |
| Min samples to split | 2 | 14 | 2 |
| Number of estimators | 80 | 200 | 10 |

For each combination of these hyperparameters, we performed 3-fold cross-validation. The combination that maximized the validation accuracy was chosen as the optimal hyperparameters. Given the optimal hyperparameters, we trained a random forests model using the entire SMOTE-balanced training data set and evaluated the performance using the holdout unbalanced testing data set.

We performed all analyses and implemented the RIS algorithm in Python.

## RESULTS

### Individual characteristics

Because our data were collected from the Women’s Health Center at VH, all individuals in our data set are female. The average age of the 103 refugees and 930 non-refugees in our dataset was not significantly different (29.6 vs. 28.4 years old, respectively, Student t-test *P* = 0.07, **Fig. 1A**). Most of the individuals live in the Phoenix metropolitan area, as seen in (**Fig. 1B-C**).

**Figure 1.**
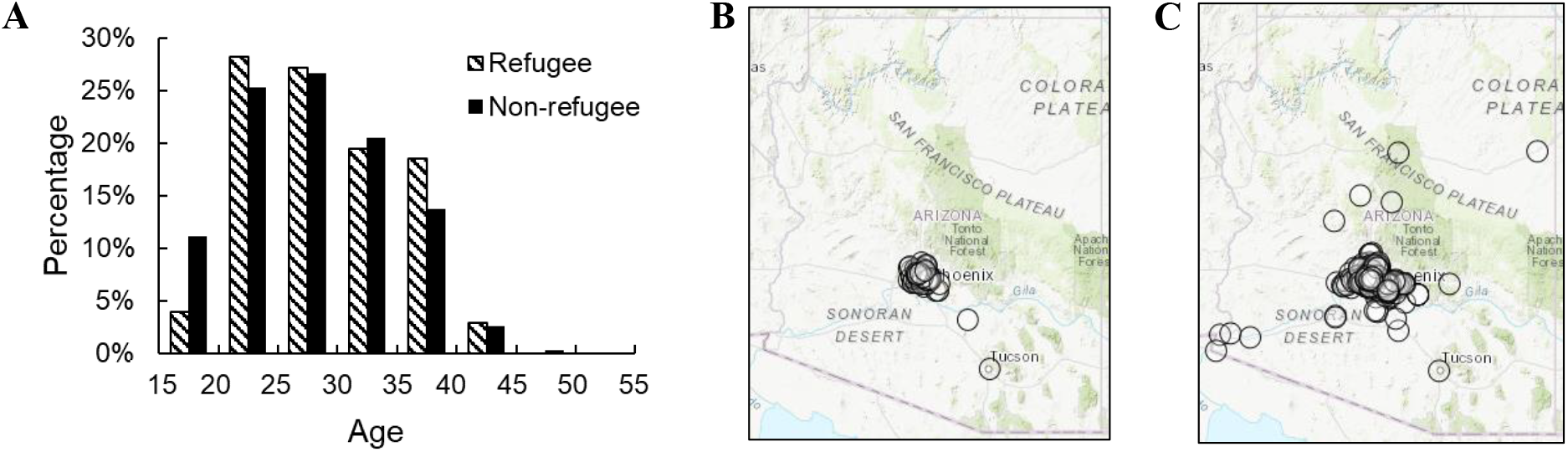
Characteristics of individuals in the curated data set. (**A**). Histograms of ages of refugees (hashed bars) and non-refugees (solid bars). (**B, C**) Geographic mapping of noise added residential addresses of refugees (**B**) and noise added non-refugees (**C**). Map of Arizona is displayed with black circles representing individual residential addresses.

### Engineered language features reflect relative frequencies in refugees and non-refugees

The ranked language list derived from Arizona state-level statistics and the CIA’s World Factbook contained 278 languages spoken in 96 countries. Spanish received the top ranking of 1. Thirteen languages tied and received the bottom ranking of 266, including Akyem, Asante, Boron, Dagarte, Dangme, Fante, Ga, German, Kokomba, Latvian, Lithuanian, Polish, and Silesian.

Although this list did not use individual-level information, the rankings correctly reflected language frequencies in the curated data set. Specifically, eight languages are spoken by at least 5% of the refugees in the curated data set, including Swahili, Kinyarwanda, English, Arabic, Burmese, Kirundi, Somali, and French. Five of these languages received rankings better than 7, and the remaining three languages received rankings better than 45 (top 16 percentile, **Table 2**).

**Table 2.** Languages spoken by >5% of refugees in the curated data and top languages in the rank list.

|  | Language | Frequency in the curated data set |  | Ranking in the language list |
| --- | --- | --- | --- | --- |
|  |  | Among refugees | Among non-refugees |  |
| Sorted by frequency in curated refugee data | Swahili | 18% | 0.2% | 45 |
|  | Kinyarwanda | 18% | 0.3% | 34 |
|  | English | 15% | 57% | 7 |
|  | Arabic | 10% | 0.3% | 2 |
|  | Burmese | 8% | 0.3% | 3 |
|  | Kirundi | 7% | 0% | 28 |
|  | Somali | 7% | 0.2% | 5 |
|  | French | 5% | 0.2% | 6 |
| Sorted by ranking in the engineered language list | Spanish | 0% | 41% | 1 |
|  | Arabic | 10% | 0.3% | 2 |
|  | Burmese | 8% | 0.3% | 3 |
|  | Kurdish | 0% | 0% | 4 |
|  | Somali | 7% | 0.2% | 5 |
|  | French | 5% | 0.2% | 6 |
|  | English | 15% | 57% | 7 |
|  | Turkmen | 0% | 0% | 8 |
|  | Dari | 3% | 0.1% | 9 |
|  | Lingala | 0% | 0% | 10 |

The top-ranking languages in the engineered list also included languages absent from our curated data set (**Table 2**). This accounts for languages that may be commonly spoken by refugees but not captured in our data set due to its limited size, allowing our model to accommodate unseen languages for better generalizability. For example, Kurdish (ranked 4^th^) is commonly spoken in Iraq, the country with the highest number of refugees resettled in Arizona since 2000^[3]^. Turkmen (ranked 8^th^) is commonly spoken in Turkmenistan, Iran, and Afghanistan from where more than 4,000 refugees fled and resettled in Arizona since 2000. We also observed a small number of languages present in our curated data set but not in the CIA World Factbook, such as Karen and Rohingya. To account for these undocumented languages, we assigned them a ranking of 50.

### Engineered location features show differences in refugee and non-refugee residential areas

Our hierarchical clustering analysis identified 19 geographic clusters of refugee addresses and 9 clusters of non-refugee addresses. Almost all refugee clusters were in the Central Phoenix metropolitan area, with only one exception in Tucson, a major refugee service site in Arizona (**Fig. 2A, 2C**). Contrarily, non-refugee clusters are scattered across Arizona (**Fig. 2B, 2D**). Although many non-refugees also live in Phoenix, the geographic clusters spread into the peripheral suburbs. These clustering patterns highlighted geographic differences between refugee and non-refugee communities.

**Figure 2.**
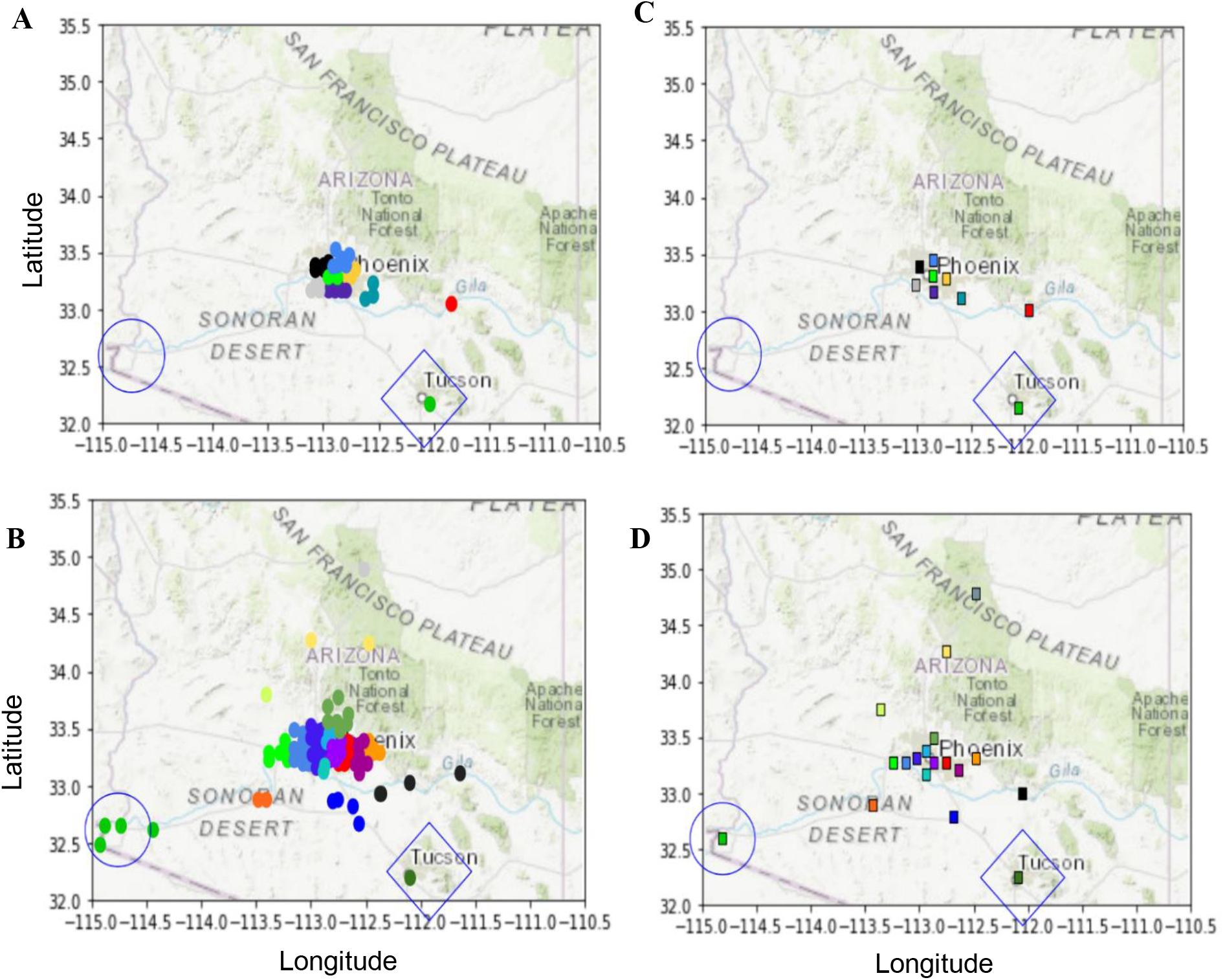
Clustering of residential addresses. (**A, B**) Geographic maps of Arizona show clusters of refugee (A) and non-refugee (**B**) addresses. Each colored dot represents an individual noise added residential address. Colors indicate different clusters. Circles and diamonds mark Yuma and Tucson, respectively. (**C, D**) Geographic maps show centroids of clusters of refugee (**C**) and non-refugee (**D**) addresses. Each colored box represents a centroid.

The geographic patterns provided information complementary to language features. For example, both Yuma and Tucson are highly populated cities with >40% of the population speaking Spanish[15]. However, we observed only a non-refugee cluster in Yuma, while Tucson harbored both refugee and non-refugee clusters. This is consistent with the fact that Yuma is not a refugee service site.

### RIS has high classification accuracy

The random forest model took 31 input features (age, language rank, and 29 location distances). The best hyperparameters determined via the grid search included maximum features = 24, minimum samples per leaf = 20, minimum samples to split = 2, and number of estimators (trees) = 40.

The random forest model showed high classification performance. Evaluated using holdout testing data, this model achieved the overall accuracy of 0.97, specificity of 0.98, sensitivity of 0.85, positive predictive value of 0.88, and negative predictive value of 0.98. Cross-validation performance was very similar to holdout testing results (**Table 3, Fig. 3A-B**). The receiver operating characteristic (ROC) curve had an area under the curve (AUC) value of 0.98 (**Fig. 3C**).

**Table 3.** Model performance metrics.

| Metric | Cross-Validation | Holdout Testing |
| --- | --- | --- |
| Accuracy | 0.96 | 0.97 |
| Specificity | 0.98 | 0.99 |
| Sensitivity (Recall) | 0.86 | 0.85 |
| Positive Predictive Value (Precision) | 0.79 | 0.88 |
| Negative Predictive Value | 0.98 | 0.98 |

**Figure 3.**
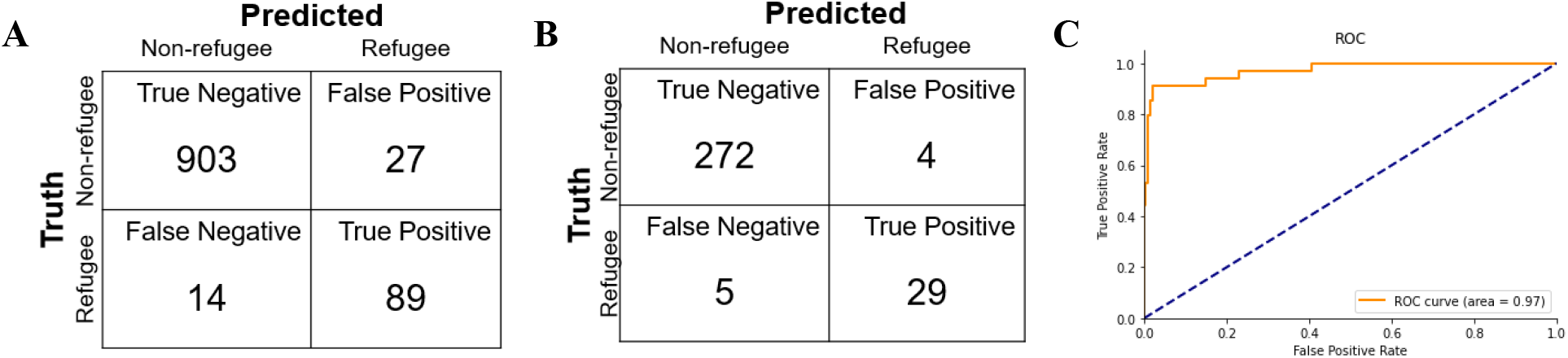
Model performance. (**A**) Confusion matrix derived from cross-validations using the training data set. (**B**) Confusion matrix derived from holdout testing data set. (**C**) ROC curve.

## DISCUSSION

RIS is the first method that uses machine-learning to identify refugees, a vulnerable population suffering severe health disparities. RIS has three favorable characteristics that make it a valuable tool to create refugee registries. First, it requires only basic information of an individual (age, primary language, and residential address) that is readily available in EMR, EHR, and insurance claims. Second, it achieves a high classification accuracy of 0.97 Third, it can be easily trained using a medium-size curated data set (1,033 individuals in our study) and state-level refugee statistics. Therefore, the computational framework is adaptable to a variety of healthcare databases in different states and organizations to build refugee registries. Due to the highly dynamic resettlement programs and expanding refugee communities, frequent updates of refugee registries are needed. In these cases, RIS can be easily re-trained, allowing it to incorporate newly emerged patterns.

In the development of RIS, we performed robust feature engineering to convert two qualitative features, namely primary language and residential address, into 29 quantitative features. This process demonstrated the power of combining individual-level information with state-level and world-level summary statistics, which allowed us to account for unseen values in the limited curated data set, improving the generalizability of RIS.

RIS classifications are not 100% accurate. To better understand the errors, we examined the misclassified individuals. We found that many refugees with English as their primary language were misclassified as non-refugees, which contributed to the entire set of false negative predictions. On the other hand, all the four false positive predictions involved individuals whose primary language is not English but ranks among the top 100 in the engineered language list. These weaknesses can potentially be addressed by incorporating additional predictors to our model, such as an individual’s citizenship and visits to refugee clinics. We refrained from adding these predictors because they are not commonly recorded in healthcare databases. However, we plan to build a nested model for individuals who provide such extra information.

Due to EMR access control, our curated data set consists of only adult female individuals visiting the Women’s Health Center. The focus on this cohort in a single site limits the generalizability of RIS. However, the computational framework is adaptable to address similar challenges in other States. In our future work, we will encompass other primary care fields of medicine including Family Medicine and Pediatrics, to enhance the power and utility of a refugee registry. We also plan to scale RIS to state-level refugee health data through Arizona’s Medicaid agency (Arizona Health Care Cost Containment System) and the state Department of Economic Security Refugee Resettlement Program.

## CONCLUSION

Refugees face special health concerns as well as disadvantaged access to and navigation of healthcare services^[16]^. RIS allows researchers, public health organizations, and healthcare practitioners to identify refugees in their databases with high accuracy and minimal manual effort. It is a critical first step to highlight refugees who are in urgent need of tailored care and support, in order to improve both their access to care and their long-term health and well-being.

## Data Availability

Data are available upon request.

## CONTRIBUTION STATEMENTS

L.L., C.E.J, and C.B. conceived of the presented idea. L.L., M.M. and V.N. developed the method. All authors contributed to the manuscript.

